# CETP and SGLT2 inhibitor combination therapy improves glycemic control

**DOI:** 10.1101/2023.06.13.23291357

**Authors:** Bohdan B. Khomtchouk, Patrick Sun, Marc Ditmarsch, John J.P. Kastelein, Michael H. Davidson

**Author notes:** Correspondence to: Bohdan Khomtchouk, PhD, 535 W. Michigan Street, Informatics & Communications Technology Complex (IT) 477, Indianapolis, IN 46202 USA.

## Abstract

**Importance:** Cholesteryl ester transfer protein (CETP) inhibition has been associated with decreased risk of new-onset diabetes in past clinical trials exploring their efficacy in cardiovascular disease and can potentially be repurposed to treat metabolic disease. Notably, as an oral drug it can potentially be used to supplement existing oral drugs such as sodium-glucose cotransporter 2 (SGLT2) inhibitors before patients are required to take injectable drugs such as insulin.

**Objective:** To identify whether CETP inhibitors could be used as an oral add-on to SGLT2 inhibition to improve glycemic control.

**Design, Setting, and Participants:** 2×2 factorial Mendelian Randomization (MR) is performed on the general population of UK Biobank participants with European ancestry.

**Exposures:** Previously constructed genetic scores for CETP and SGLT2 function are combined in a 2×2 factorial framework to characterize the associations between joint CETP and SGLT2 inhibition compared to either alone.

**Main Outcomes and Measures:** Glycated hemoglobin and type-2 diabetes incidence.

**Results:** Data on 233,765 UK Biobank participants suggests that individuals with genetic inhibition of both CETP and SGLT2 have significantly lower glycated hemoglobin levels (mmol/mol) than control (Effect size: −0.136; 95% CI: −0.190 to −0.081; p-value: 1.09E-06), SGLT2 inhibition alone (Effect size: −0.082; 95% CI: −0.140 to −0.024; p-value: 0.00558), and CETP inhibition alone (Effect size: −0.08479; 95% CI: −0.136 to −0.033; p-value: 0.00118).

Furthermore, joint CETP and SGLT2 inhibition is associated with decreased incidence of diabetes (log-odds ratio) compared to control (Effect size: −0.068; 95% CI: −0.115 to −0.021; p-value: 4.44E-03) and SGLT2 inhibition alone (Effect size: −0.062; 95% CI: −0.112 to −0.012; p-value: 0.0149).

**Conclusions and Relevance:** Our results suggest that CETP and SGLT2 inhibitor therapy may improve glycemic control over SGLT2 inhibitors alone. Future clinical trials can explore whether CETP inhibitors can be repurposed to treat metabolic disease and provide an oral therapeutic option to benefit high-risk patients before escalation to injectable drugs such as insulin or glucagon-like peptide 1 (GLP1) receptor agonists.

**Key Points:** 

**Question:** Does combined genetic CETP and SGLT2 inhibition confer decreased glycated hemoglobin or diabetes incidence compared to SGLT2 inhibition alone?

**Findings:** In this cohort study, a 2×2 factorial Mendelian randomization analysis on the UK Biobank reveals that combined genetic CETP and SGLT2 inhibition is associated with decreased glycated hemoglobin and diabetes risk compared to both control and SGLT2 inhibition alone.

**Meaning:** Our findings suggest that CETP inhibitors, which are currently in clinical trials to treat cardiovascular disease, can be repurposed to treat metabolic disease in a combination therapy approach with SGLT2 inhibitors.

## Introduction

Metabolic disease has some of the highest medical burden in the world, with nearly 35% of people (over 100 million patients) afflicted with some form of metabolic syndrome in the United States alone, costing our healthcare system >$250 billion annually^1^. Despite the extensive amount of research performed on developing treatments for metabolic disease, there is an unmet need for understanding synergies between existing treatment when used in combination therapies, which has been a major driving force behind improved outcomes in other disease areas such as oncology. Furthermore, the growing understanding of an intersection between cardiovascular and metabolic disease represents a rich knowledge base from which combination therapies using treatments from these two intersecting disease verticals can be used to develop innovative and effective therapies to achieve better outcomes. Here, we investigate whether combining the hyperlipidemia drug class of cholesteryl ester transfer protein (CETP) inhibitors with the existing hyperglycemia treatment in sodium glucose transporter 2 (SGLT2) inhibitors is an effective strategy to improve glycemic control. SGLT2 inhibitors are a novel class of oral antidiabetic drugs that reduce glucose toxicity by stimulating its excretion into urine and inhibiting its reabsorption in the kidneys^2, 3^. Because this mechanism of action is independent of insulin secretion or action, SGLT2i can be used in combination with other therapies to improve outcomes for patients afflicted with type II diabetes mellitus (T2DM)^4^. Circulating levels of plasma CETP reduce pancreatic β-cell insulin secretion by disrupting cholesterol homeostasis through accumulation of free cholesterol, which causes β-cell lipotoxicity and dysfunction that induces T2DM^5^. Therefore, CETP inhibition is one promising approach to ameliorating β-cell function in T2DM by decreasing islet cholesterol accumulation and inflammation^5^. Since CETP inhibitors (CETPi) are known to increase HDL concentrations, which are the predominant acceptors of cell cholesterol and have been reported to inhibit β-cell apoptosis and promote β-cell survival, it follows that this therapeutic strategy may be important for maintaining normal β-cell function and insulin secretion^6^. Indeed, previous meta-analyses of existing randomized controlled clinical trials of CETPi therapies have shown that CETP inhibitors significantly reduce the incidence of new onset diabetes while improving glucose homeostasis and metabolism^7^. Therefore, in this study, we speculated that synergistic effects might be jointly induced by a combination of SGLT2i and CETPi therapy. We computationally tested this hypothesis through the design of a new 2×2 factorial Mendelian randomization study comprised of 233,765 individuals from the UK Biobank. Our study builds upon and improves on prior work exploring the efficacy of combination therapies in smaller populations that focused on other clinical indications and drugs (e.g., previous efforts to genetically mimic the effects of ezetimibe and statins in 108,376 people with coronary heart disease^8^).

## Methods

### Construction of genetic scores

To construct a genetic score that computationally mimics the biological effects of CETP inhibitors, we build a score of 4 single-nucleotide polymorphisms (SNPs) in the CETP gene region that are strongly correlated with HDL. The genetic score uses all genotyped SNPs in the UK Biobank that were included in a CETP score described in a prior study^9, 10^. A higher CETP genetic score mimics a greater degree of pharmaceutical CETP inhibition (Figure 1A,B). SNPs are filtered for inclusion such that they are all approximately in linkage disequilibrium (LD) with r^2<0.3 (Supplementary Table 4 and 5).

**Figure 1:**
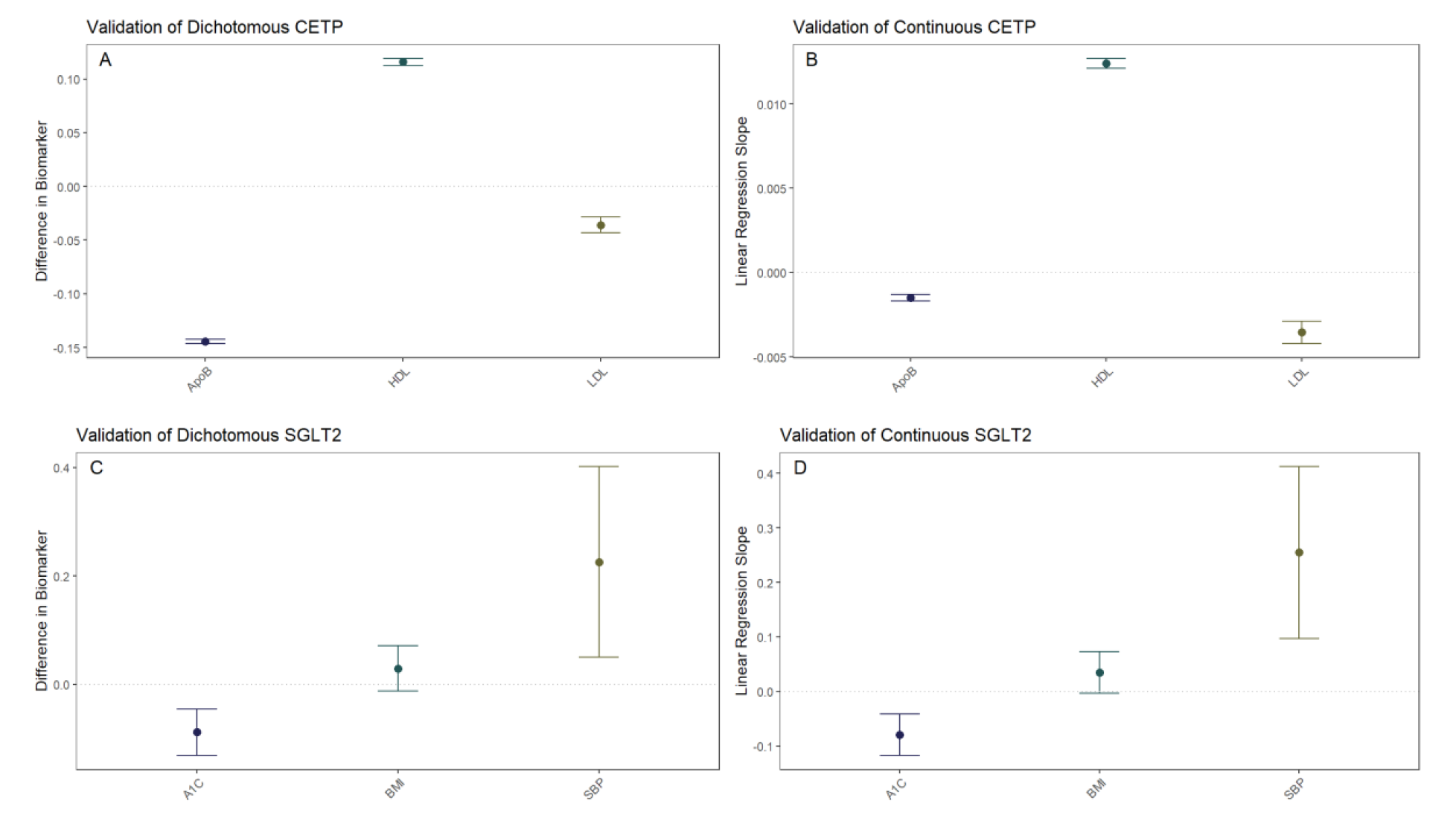
**Construction of Genetic Scores** Both dichotomous **A)** and continuous **B)** CETP scores are associated with decreased ApoB, increased HDL, and decreased LDL. Point estimates and 95% confidence intervals are plotted for the difference between the high-CETP score group. Both dichotomous **C)** and continuous **D)** SGLT2 scores are associated with decreased hemoglobin A1c and increased systolic blood pressure, with increased BMI trending towards significance.

To construct a genetic score that mimics the effects of SGLT2 inhibitors, we build a score of 2 single-nucleotide polymorphisms (SNPs) in the SGLT2 gene region that are strongly correlated with SGLT2 expression. The genetic score uses all genotyped SNPs in the UK Biobank genotyping information that were included in the SGLT2 score described in Katzmann et al., 2021^9, 11^. A higher SGLT2 genetic score mimics a greater degree of pharmaceutical SGLT2 inhibition (Figure 1C,D).

### Instrumental variable data analysis

We perform a 2×2 factorial Mendelian Randomization (MR) analysis using the UK Biobank with a focus on CETP and SGLT2-relevant datasets, as outlined in Figure 2A and Figure 2C. We include all individuals in the UK Biobank that have genotyping information containing all SNPs needed to construct the genetic scores, as well as all biomarker values. Furthermore, only white British individuals are included in the analysis to control for population stratification bias. This is performed because the SGLT2 score from Katzman et al., 2021 is validated only in the white population of the UK Biobank since the eQTLs from which they were constructed were identified in a primarily white population^11, 12^. In total, 233,765 individuals fulfil our study’s inclusion criteria. The final study population is then analyzed with the 2×2 factorial MR methodology detailed in Figure 2B.

**Figure 2:**
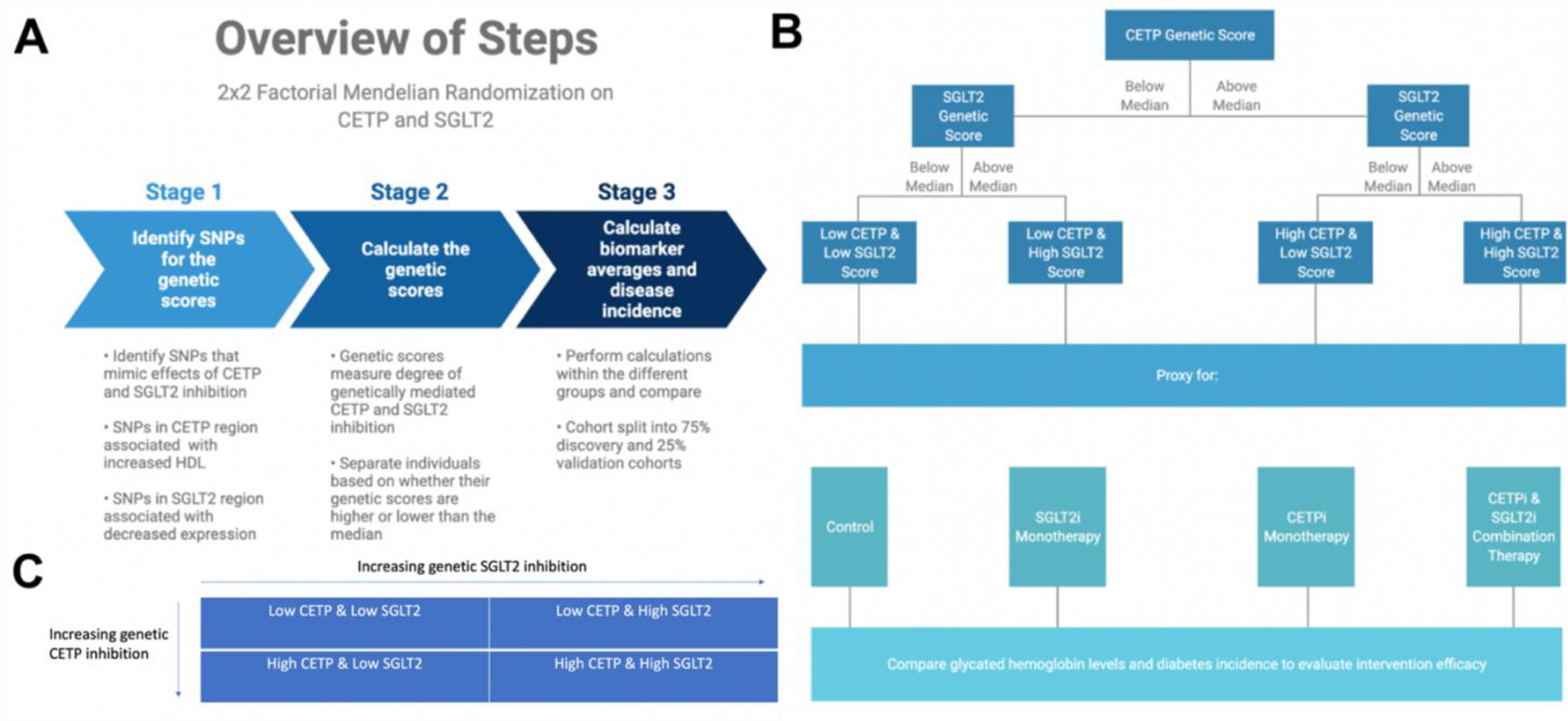
**Graphical abstract of methods overview. A)** Overview of computational workflow for the 2×2 factorial Mendelian Randomization (MR) split into 3 steps: identifying SNPs for the genetic scores that mimic effects of CETP and SGLT2 inhibition, calculating the genetic scores, calculating biomarker averages and disease incidence for each of the four groups. **B)** Description of allocating individuals into one of the four groups: Low CETP and low SGLT2 score, low CETP and high SGLT2 score, high CETP and low SGLT2 score, and high CETP and high SGLT2 score. The same 2×2 factorial MR methods were conducted with each group. **C)** Once genetic scores were obtained and individuals separated into one of the four aforementioned groups, a 2×2 factorial MR analysis was performed using UK Biobank with a focus on CETP and SGLT2-relevant datasets comprising 233,765 unique individuals.

Individuals are separated into two groups based on whether their CETP genetic score is greater than or less than the median CETP score. In each group, individuals are then separated into two additional groups based on whether their SGLT2 genetic score is greater than or less than the median SGLT2 genetic score. In total, four groups are formed. Then, the mean age, sex, HDL, LDL, TG, ApoB, weight, systolic blood pressure (SBP), glycated hemoglobin, and diabetes incidence rates are recorded for each of the four groups in both the discovery and replication cohorts. Differences in quantitative variables between groups are evaluated using linear regression and differences in diabetes incidence are calculated using logistic regression. We conduct analysis controlling for body mass index (BMI) and systolic blood pressure (SBP) using linear regression for glycated hemoglobin and logistic regression for diabetes, though analysis excluding SBP and BMI as covariates is also conducted. The covariates of BMI and SBP are included in the regression models because the SGLT2 genetic score used to mimic SGLT2 inhibition is associated with increased SBP and BMI, which is the opposite direction of effect that pharmaceutical SGLT2 inhibition has on these variables. BMI and SBP are included as covariates to ensure that the difference in glycated hemoglobin observed between groups is not due to these associations. Diabetes incidence is defined by International Classification of Diseases (ICD) codes retrieved from the electronic medical health records database associated with the UK Biobank. All individuals included in this analysis have available health records. Non-additive effects between genetic CETP and SGLT2 scores are detected through linear regressions with CETP, SGLT2, and their product (non-additive term) against glycated hemoglobin and logistic regression against diabetes incidence. Interaction is detected as a significant p-value in the interaction term of the regression.

### Statistical Analysis

All analysis is performed in the R programming language, v4.1.0^13^. All visualization is performed using the ggplot2 package in R^14^. All hypothesis tests are two-sided and use a statistical significance level of 0.05.

### Study/Ethics Approval

All participants gave written informed consent prior to data collection. UK Biobank has full ethical approval from the NHS National Research Ethics Service (16/NW/0274). All methods were carried out in accordance with relevant guidelines and regulations. UK Biobank data is available to researchers upon request (https://www.ukbiobank.ac.uk/enable-your-research).

## Results

### Confirming Genetic Score Function

Pharmaceutical CETP inhibition is known to increase HDL and decrease LDL along with ApoB^15,16,17,18,19^. Thus, we validated whether our CETP genetic score behaved similarly to pharmaceutical CETP inhibition by seeing if elevated CETP genetic scores, corresponding to more CETP inhibition, exhibited these same relationships. This validation was performed through a dichotomous CETP score, where participants are split into high and low groups and differences in lipid biomarkers are observed, as well as through a continuous CETP score, where the score itself is regressed against HDL, LDL, and ApoB. We find both scores are strongly associated with increased HDL, decreased LDL, and decreased ApoB with p-values less than 2×10^-16 for all comparisons (Figure 1A, 1B; Supplementary Table 1). Based on these results, our CETP score is likely a reasonable proxy for pharmaceutical CETP inhibition.

Pharmaceutical SGLT2 inhibition decreases glycated hemoglobin, systolic blood pressure, and body mass index (BMI), so we validate the function of our SGLT2 genetic score based on associations with these biomarkers^20,21,22,23^. We perform the same analysis with dichotomous and continuous scores as detailed above to validate our CETP genetic score, finding that our SGLT2 score is associated with decreased glycated hemoglobin (dichotomized: p=4.26×10^- 5; continuous: p=4.40×10^-5), but increased systolic blood pressure (dichotomized: p=0.0116; continuous: p=0.00149) with increased BMI trending towards significance (dichotomized: p=0.172; continuous: p=0.0708) (Figure 1C, 1D; Supplementary Table 1). Based on these results, we conclude that our SGLT2 score functions as expected with respect to glycated hemoglobin, but not with respect to SBP and BMI. Since the primary mechanism of action of SGLT2 inhibition is to decrease glycated hemoglobin with decreased blood pressure and BMI as secondary effects, we believe our SGLT2 score is likely a reasonable proxy for pharmaceutical SGLT2 inhibition. However, we will control for the possible confounding of the SBP and BMI associations by including them as covariates in all aspects of the following 2×2 factorial Mendelian Randomization analysis. For each comparison between groups there will be two statistical tests performed: one without SBP and BMI as covariates and one including SBP and BMI as covariates.

### Comparison of glycated hemoglobin between groups

After confirming the function of our scores and identifying SBP and BMI as possible confounders to control for, we proceed with our 2×2 factorial Mendelian Randomization analysis and split our cohort into four groups corresponding to control, SGLT2 inhibition only (SGLT2i), CETP inhibition only (CETPi), and both SGLT2 and CETP inhibition (combo therapy). We compare the levels of glycated hemoglobin between members of each group and the reference group (low CETP and low SGLT2 score). We find that both SGLT2i (p=0.0719) and CETPi (p=0.0549) are associated with decreased glycated hemoglobin relative to control, and this signal is strengthened to statistical significance (SGLT2i: p=0.0087; CETPi: p=0.044) when SBP and BMI are included as covariates (Figure 3A, 3B; Supplementary Table 2). Furthermore, when the non-control groups are compared against each other, no significant difference is found between the CETPi and SGLT2i groups (p=0.915), but the combo therapy group has significantly lower glycated hemoglobin levels relative to the SGLT2i (p=0.00558) and CETPi (p=0.00118) groups; this conclusion is recapitulated when SBP and BMI are included as covariates (CETPi vs SGLT2i: p=0.384; Combo vs SGLT2i: p=0.0459; Combo vs CETPi: p=0.00146) (Figure 3C, 3D; Supplementary Table 2). Non-additive effects between genetic CETP and SGLT2 inhibition on glycated hemoglobin are not detected, as the interaction term is not significantly associated with glycated hemoglobin (Estimate: 9.13×10^- 5; 95% CI: −0.0007 to 0.0007; p-value=0.98) (Supplement 9).

**Figure 3:**
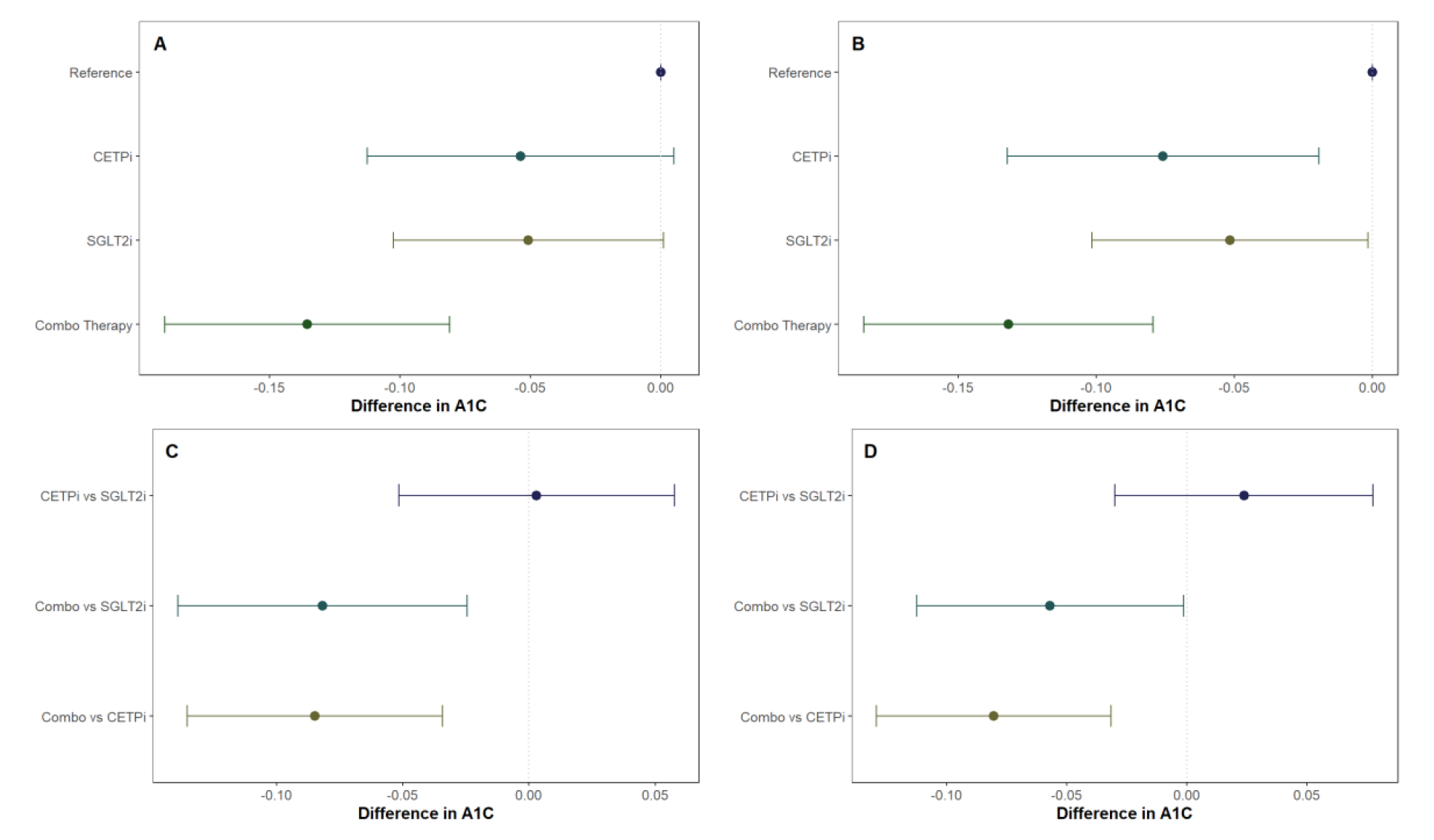
All groups in the 2×2 factorial Mendelian Randomization framework are compared against control to see if they have lower glycated hemoglobin levels. This is performed **A)** without SBP and BMI as covariates and **B)** with SBP and BMI as covariates. Non-control groups are then compared with each other **C)** without SBP and BMI as covariates and **D)** with SBP and BMI as covariates.

### Comparison of diabetes risk between groups

To determine whether joint CETP and SGLT2 inhibition has an impact on diabetes risk within each of our groups, we leverage the same analytical framework as detailed above for glycated hemoglobin but use logistic regression instead of linear regression because diabetes is a binary phenotype. We find that neither CETPi nor SGLT2i are associated with decreased diabetes risk relative to control (CETPi: p=0.806; SGLT2i: p=0.274), but combo therapy is associated with significantly decreased diabetes risk relative to control (p=4.44×10^-3) (Figure 4A; Supplementary Table 3). These findings are unchanged if SBP and BMI are included as covariates (CETPi: p=0.446; SGLT2i: p=0.233; Combo: p=3.82×10^-3) (Figure 4B; Supplementary Table 3). Comparison between groups suggests that SGLT2i and CETPi do not have significantly different incidence of diabetes (p=0.444), even though the combo therapy group is significantly associated with decreased diabetes incidence relative to SGLT2i (p=0.0149) and is trending towards significance relative to CETPi (p=0.0557) (Figure 4C; Supplementary Table 3). Replication of these findings is achieved when SBP and BMI are included as covariates (CETPi vs SGLT2i: p=0.739; Combo vs SGLT2i: p=0.0466; Combo vs CETPi: p=0.0609) (Figure 4D; Supplementary Table 3). Non-additive effects between genetic CETP and SGLT2 inhibition on diabetes risk are not detected, as the interaction term is not significantly associated with diabetes risk (Estimate: −0.00233; 95% CI: −0.008 to 0.004; p- value=0.454) (Supplement 9). A summary of all evaluated quantities between different groups in the 2×2 factorial framework is presented in Figure 5.

**Figure 4:**
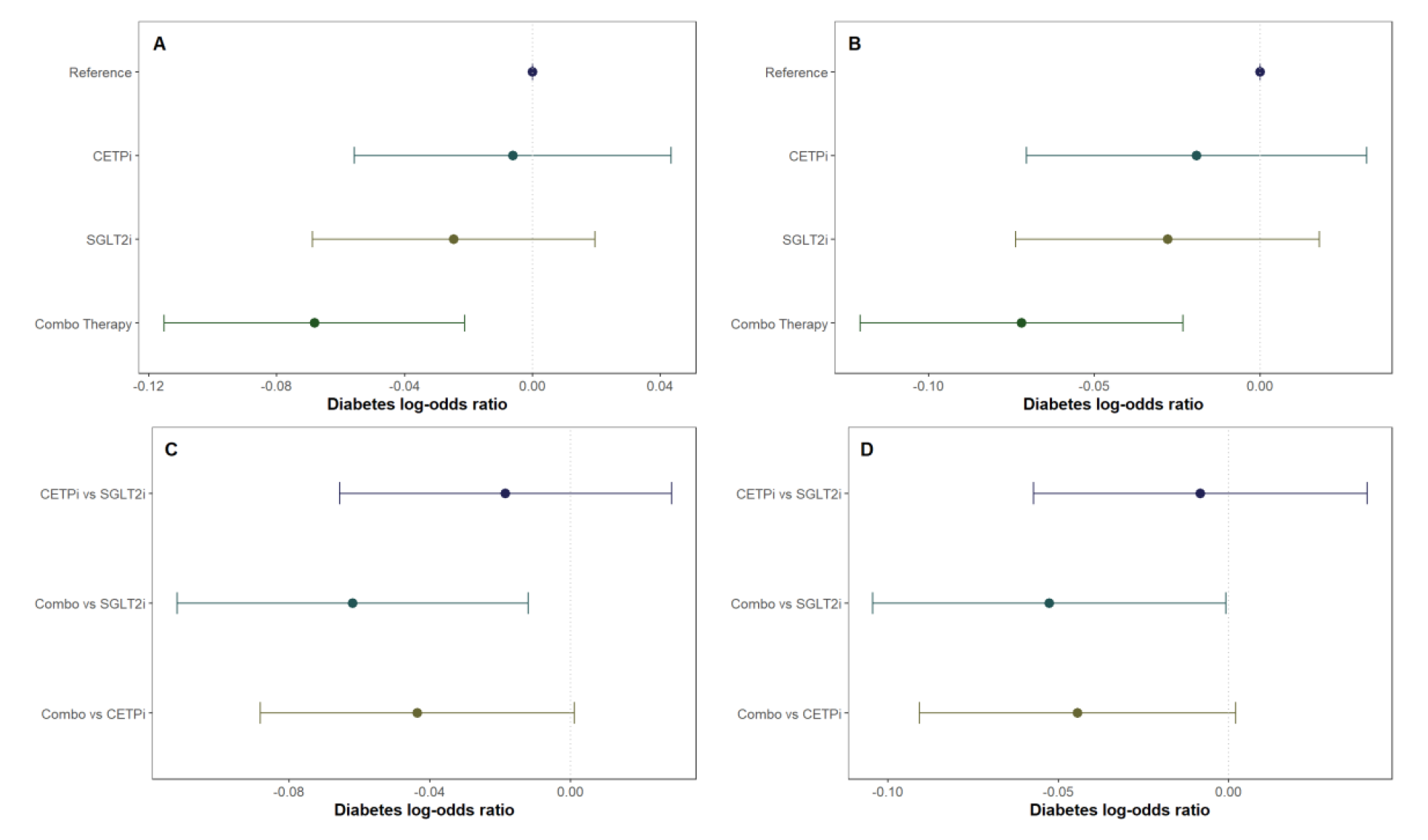
All groups in the 2×2 factorial Mendelian Randomization framework are compared against control to see if they have lower diabetes incidence. This is performed **A)** without SBP and BMI as covariates and **B)** with SBP and BMI as covariates. Non-control groups are then compared with each other **C)** without SBP and BMI as covariates and **D)** with SBP and BMI as covariates.

**Figure 5:**
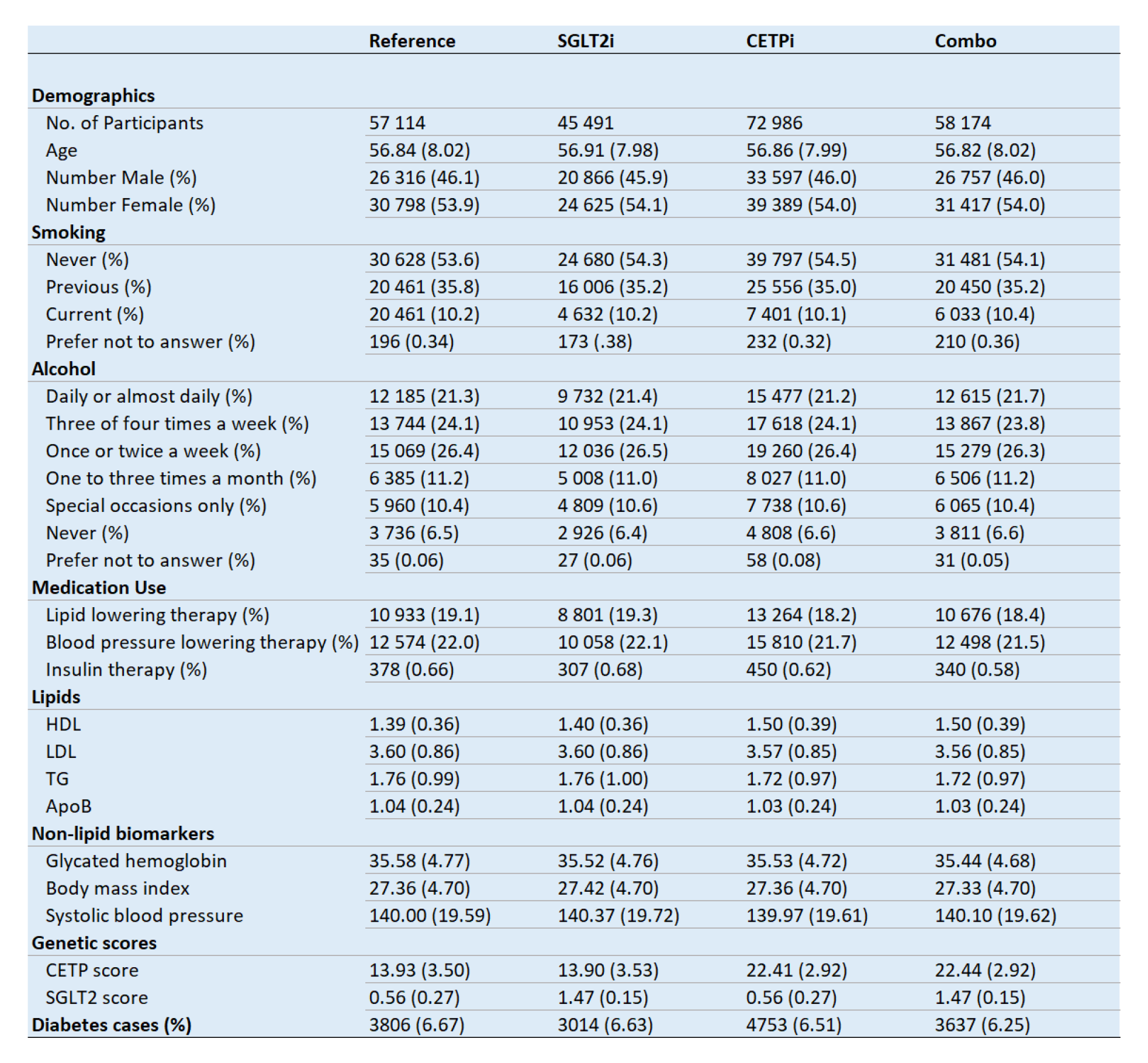
**Regression of genetic scores against biomarkers.** Demographic, biomarker, genetic score, and diabetes data differences between the different groups included in the 2×2 factorial Mendelian randomization design.

## Discussion

In this study, we have used a 2×2 factorial Mendelian Randomization framework to investigate the effects of SGLT2i and CETPi combination therapy on glycated hemoglobin levels and diabetes risk. We find that combination therapy is associated with decreased glycated hemoglobin levels compared to SGLT2i, CETPi, and control, with this conclusion robust against the inclusion of potential confounders (SBP and BMI) as covariates. The combination therapy group also had significantly lower diabetes incidence compared to both control and SGLT2i and was trending towards significance for CETPi. We detected no evidence of interactions between genetic CETP and SGLT2 inhibition on either glycated hemoglobin or diabetes risk. Taken together, these results constitute genetics-driven evidence suggesting that combination therapy with CETP and SGLT2 inhibitors confers improved protection against hyperglycemia and diabetes risk compared to SGLT2 inhibitors alone. Furthermore, the lack of interaction effect suggests that genetic CETP or SGLT2 inhibition doesn’t attenuate the effect of the other on glycated hemoglobin or diabetes risk. It is important to note that since CETP and SGLT2 inhibitors are both oral drugs, their combination therapy represents an oral therapeutic strategy for treatment-resistant diabetes prior to use of injectable drugs such as insulin or glucagon-like peptide 1 (GLP1) receptor agonists^24^. The ease of taking oral therapy over injectable drugs could lead to higher patient compliance to drive better patient outcomes as well as lower metabolic disease morbidity. Indeed, previous investigations of medication adherence in type-2 diabetes estimate adherence of oral hyperglycemia agents to be between 38% and 93%, generally higher than that of injectable hyperglycemia agents, which are between 38% and 61%^25,26,27,28,29^.

The cost of bringing a new drug to market is exceptionally high, costing over $1 billion with estimates ranging as high as $2.6 billion^30, 31^. Beyond heavy monetary investment, developing medicine to treat a disease from scratch requires a time investment of over a decade^31, 32^. Finding ways to repurpose existing drugs into promising new clinical indications represents an efficient way to decrease drug discovery cost and development time. Critically, since CETP inhibitors such as obicetrapib are currently in phase 3 clinical trials to treat dyslipidemia and coronary artery disease, its clean safety and toxicity profile makes it a good repurposing candidate for metabolic disease and combination therapies^33^. Our analysis not only provides genetics-driven support for repurposing CETP inhibitors to metabolic disease in the form of a CETP and SGLT2 inhibitor combination therapy, but also highlights the power of 2×2 factorial Mendelian Randomization as a computational framework through which combination therapies across disease verticals can be systematically discovered and mined for repurposing opportunities. Previous 2×2 factorial MR studies have identified combination therapies primarily within cardiovascular disease for PCSK9 and CETP inhibition, NPC1L1 and HMGCR inhibition, and IL-6 and PCSK9/CETP/NPC1L1 inhibition^8, 34, 35^. However, our study is the first to use 2×2 factorial MR as a way to identify phase 3 cardiovascular drugs that can be repurposed for metabolic disease in a combination therapy approach. This conclusion is currently patent-pending under application number 37726-54665.

Despite the translational impact of our findings, our study is not without its limitations. The primary shortcoming of our analysis is the fact that we did not conduct a randomized double-blind placebo-controlled clinical trial, which is the gold benchmark standard, to administer pharmacologic SGLT2 or CETP inhibitors to investigate the effect of either monotherapy. Rather, we computationally analysed the effect of lifetime decreased SGLT2 or CETP function caused be genetic variation, which could differ from the shorter-acting effects of pharmaceutical therapies. Since our approach is grounded in genetics, it also doesn’t account for the potential off-target effects of small molecule inhibitors of either SGLT2 or CETP, which can only be achieved through a randomized control trial as mentioned above. However, our analysis does provide strong genetics-driven evidence grounded in a well-established causal inference framework that can be used to inform future clinical trials of this combination therapy. Furthermore, the actual instruments may have pleiotropic effects on glycated hemoglobin or lipid parameters independent of their effect on SGLT2 or CETP function, though we have attempted to minimize this potential source of confounding by using previously validated genetic scores from the published literature. Lastly, our analysis demonstrates the effectiveness of joint CETP and SGLT2 inhibition in a white population within the UK Biobank, but the strength and generalizability of our analysis would be increased by are more ethnically diverse data cohort, either by including all individuals in the UK Biobank or by using a more heterogeneous cohort such as the NIH *All of Us* Research Program^36^.

## Conclusion

Genetically downregulated SGLT2 signaling and genetically lowered CETP activity are associated with additively lower lifetime risk of metabolic outcomes by way of significantly decreased glycated hemoglobin levels and lower incidence of type-2 diabetes. Future clinical trials should explore the protective effects of combining SGLT2 inhibitors and CETP- lowering treatments in the context of metabolic disease prevention.

## Contributions

BBK, MD, JJP, MHD designed the research studies. BBK, PS acquired data. BBK, PS analyzed data and helped with data interpretation and figure generation. BBK wrote the original draft of the manuscript. All authors reviewed and edited the manuscript.

## Data Availability

All data produced are available online at: https://www.ukbiobank.ac.uk/enable-your-research

## Acknowledgement

BBK acknowledges funding support from Indiana University and NIH R01DK132090.

